# Pharmacokinetics of anti-tuberculosis drugs in multidrug resistant tuberculosis patients in India

**DOI:** 10.1101/2020.05.26.20111534

**Authors:** Kumar AK Hemanth, PL Natarajan, T Kannan, R Sridhar, S Kumar, Kumar V Vinod, NS Gomathi, T Bharathiraja, V Sudha, S Balaji, S Rameshkumar, Dina Nair, SP Tripathy, R Geetha

## Abstract

Programmatic Management of multidrug resistant tuberculosis (MDR TB) services were introduced in the Indian TB control programme in 2007. A pharmacokinetic (PK) study of drugs used to treat MDR TB, namely levofloxacin (LFX), ethionamide (ETH), cycloserine (CS), pyrazinamide (PZA), moxifloxacin (MFX) and isoniazid (INH) was undertaken in adult MDR TB patients treated according to the prevailing guidelines in India. Factors influencing drug PK and end-of-intensive phase (IP) status were also determined. We recruited 350 MDR TB patients receiving anti-TB treatment (ATT) in the Indian Government programme in south India. At steady state, serial blood samples were collected, after supervised drug administration. Status at end of IP was noted from the programme records. Of the 303 patients for whom end-of-IP status was known, 214 were culture negative (responders), while 45 patients were either culture positive or required change of regimen or had died before completion of IP (non-responders). The median C_max_ (2.0 vs 2.9μg/ml; p = 0.005) and AUC_0-12_ (12.2 vs 17.0μg/ml.h; p = 0.002) of ETH were significantly lower in non-responders than responders at IP. In multivariate logistic regression analysis, after excluding defaulters and adjusting for confounders, AUC_0-12_ of ETH significantly influenced end-of-IP status (aOR - 1.065; 95% CI: 1.001 - 1.134; p = 0.047). Drug doses used currently in the programme produced optimal drug concentrations in majority of patients. ETH played a major role in the MDR TB combination regimen and was a key determinant of end-of-IP status.

---

The burden of multidrug-resistant (MDR) tuberculosis (TB) is of major interest and concern at global, regional and country levels. In 2018, there were approximately half a million (range 417000–556000) new cases of rifampicin-resistant (RR) TB, of which 78% had MDR TB (1). India accounts for 27% of the total MDR TB cases worldwide, which is the highest for any country. This is followed by China (14%) and the Russian federation (9%).

The National TB Elimination Programme (NTEP) in India had introduced Programmatic Management of MDR TB (PMDT) services in 2007, and a complete geographic coverage was achieved in 2013 (2). Under this programme, MDR TB patients were treated with six drugs which included an aminoglycoside and a fluoroquinolone for a total period of 24 months. The intensive phase (IP) of treatment was for 6 months comprising of kanamycin (Km), levofloxacin (LFX), ethionamide (ETH), cycloserine (CS), pyrazinamide (PZA) and ethambutol (EMB) daily, followed by the continuation phase of treatment for the remaining 18 months with LFX, Eth, CS and EMB daily. If the 4^th^ month culture for *M. tuberculosis* was positive, the intensive phase was extended up to 9 months. Subsequently, in 2018, the regimens were revised and the duration of treatment was shorter for a period of 9 months. In the revised regimen, the initial IP was for 4 months, during which patients received Km, moxifloxacin (MFX), high dose isoniazid (INH), PZA, clofazimine (CFZ), Eth and EMB daily. This was followed by the continuation phase for the remaining 5 months, during which patients received MFX, CFZ, PZA and EMB daily. In both regimens, drug doses were based on body weight and were available in four weight bands, namely, 16 - 29 kg, 30 - 45 kg, 46 - 70 kg and > 70 kg (Table 1).

**Table 1:** Drug doses and weight bands followed in the PMDT guidelines.

| INTENSIVE PHASE (6 MONTHS) |  |  |  |  |
| --- | --- | --- | --- | --- |
| DRUGS | 16 - 29 kg | 30 - 45 kg | 46 - 70 kg | ABOVE 70 kg |
| LFX | 250 | 750 | 1000 | 1000 |
| ETH | 375 | 500 | 750 | 1000 |
| CS | 250 | 500 | 750 | 1000 |
| EMB | 400 | 800 | 1200 | 1600 |
| PZA | 750 | 1250 | 1750 | 2000 |
| KM | 500 | 750 | 750 | 1000 |
| CONTINUATION PHASE (18 MONTHS) |  |  |  |  |
| DRUGS | 16 - 29 kg | 30 - 45 kg | 46 - 70 kg | ABOVE 70 kg |
| LFX | 250 | 750 | 1000 | 1000 |
| ETH | 375 | 500 | 750 | 1000 |
| CS | 250 | 500 | 750 | 1000 |
| EMB | 400 | 800 | 1200 | 1600 |
| MDR SHORTER REGIMEN |  |  |  |  |
| INTENSIVE PHASE (4 - 6 MONTHS) |  |  |  |  |
| DRUGS | 16 - 29 kg | 30 - 45 kg | 46 - 70 kg | ABOVE 70 kg |
| HIGH DOSE MFX | 400 | 600 | 800 | 800 |
| HIGH DOSE INH | 300 | 600 | 900 | 900 |
| PZA | 750 | 1250 | 1750 | 2000 |
| ETH | 375 | 500 | 750 | 1000 |
| CFZ | 50 | 100 | 100 | 200 |
| EMB | 400 | 800 | 1200 | 1600 |
| KM | 500 | 750 | 750 | 1000 |
| CONTINUATION PHASE ( 5 MONTHS) |  |  |  |  |
| DRUGS | 16 - 29 kg | 30 - 45 kg | 46 - 70 kg | ABOVE 70 kg |
| MFX | 400 | 600 | 800 | 800 |
| CFZ | 50 | 100 | 100 | 200 |
| PZA | 750 | 1250 | 1750 | 2000 |
| EMB | 400 | 800 | 1200 | 1600 |

LFX - Levofloxacin; ETH - Ethionamide; CS - Cycloserine; EMB - Ethambutol; PZA - Pyrazinamide; KM - Kanamycin

Cure rates observed among MDR TB patients from different studies varied from 31% to 75%, the treatment regimens being different in these studies. A study from south India on the management of MDR TB, reported a cure rate of 38%, with failure (25%), default (24%) and death (12%) (3). The reasons for development of MDR TB could be due to microbial, clinical or programmatic issues. Clinical characteristics of patients have also been recognized wherein appropriately administered drug doses may not achieve necessary drug levels to deal with all populations of mycobacteria. Maintaining therapeutic drug concentrations in blood is an important requisite to achieve satisfactory treatment outcome. The National Jewish Medical and Research Centre at Denver, CO, USA recommends measurement of serum concentrations of second-line anti-TB drugs early in the course of treatment, so that poor drug absorption can be dealt with, in a timely manner by optimizing drug doses (4).

A few studies have reported on the pharmacokinetics (PK) of second-line anti-TB drugs. A population PK study in 14 MDR TB patients from Korea has been reported (5). Another study from Tanzania has examined plasma activity of certain second-line anti-TB drugs in 25 patients with MDR TB (6). Park and others from the Republic of Korea have described the PK of second-line anti-TB medications in healthy volunteers (7). However, there is a paucity of PK data of second-line anti-TB drugs in MDR TB patients from India. We undertook a prospective study to determine the PK of certain anti-TB drugs (LFX, MFX, Eth, CS, PZA, INH) in adult MDR TB patients in south India treated according to the prevailing NTEP guidelines. We also examined factors that influenced drug PK and culture status at end of IP.

## Methods

### Patients

A prospective study was undertaken in adult patients with MDR TB receiving anti-TB treatment (ATT) at the Government Hospital for Thoracic Medicine, Chennai, India during June 2016 to September 2019. All the patients were bacteriologically confirmed to have MDR TB based on drug susceptibility tests. Diagnosis and treatment were in accordance with the NTEP guidelines (2 - 3 of protocol). All the patients received drugs from the NTEP under direct supervision.

Patients meeting the following study criteria were recruited: (i) aged 18 years or above (ii) body weight > 30kg (iii) minimum seven doses of ATT drugs (iv) not very sick or moribund and (v) willing to participate and give informed written consent. A structured questionnaire was used to collect patient details. The study was approved by the Institutional Ethics Committee.

### Study procedures

The PK study was conducted at the Government Hospital of Thoracic Medicine, Chennai, India after patients have had at least two weeks of treatment. Eligible patients were requested to get admitted to the hospital ward at least a day prior to start of the study. On the day of the study, a sample of blood (2 ml) was collected (pre-dosing). The prescribed anti-TB drugs were administered to the patients under supervision. Thereafter, serial blood samples were collected at 1,2, 4, 6, 8 and 12 hours after drug administration. Those with a known history of type 2 diabetes mellitus (DM), with or without random blood glucose ≥ 200mg/dl on the study day was considered diabetic. The patients’ body mass index (BMI) was calculated from their height and body weight. A BMI below 18.5 was considered malnourished.

### Drug estimations

Plasma concentrations of LFX, MFX, ETH, CS, INH and PZA were estimated by High Performance Liquid Chromatography (HPLC) (Shimadzu Corporation, Kyoto, Japan) according to validated methods described elsewhere. In brief, the method to estimate LFX and MFX involved deproteinisation of the sample with perchloric acid and analysis of the supernatant using a reversed-phase C18 column (150mm) using fluorescence detector set at an excitation wavelength of 290 nm and an emission wavelength of 460 nm. The mobile phase consisted of a mixture of phosphate buffer and acetonitrile. The retention times of LFX and MFX were 1.8 and 4.6 minutes respectively (8, 9).

Plasma INH and PZA were estimated simultaneously by extraction using para-hydrobenzaldehyde and trifluoro acetic acid. Analysis was performed using a C_8_ column at 267nm. The mobile phase consisted of water: methanol containing perchloric acid and tetrabutyl n-ammonium hydroxide. The retention times of PZA and INH were 3 and 5.5 minutes respectively (10).

The method for estimation of CS involved extraction of the drug using solid phase extraction cartridges. The analytical column was Atlantis T3 and the mobile phase was a mixture of phosphate buffer, acetonitrile and isopropyl alcohol. The retention time of CS was 4.8 minutes (11).

The method for estimation of ETH involved deproteinisation of the sample with perchloric acid and analysis of the supernatant using a reversed-phase CN column (150mm) and UV detector set at 267 nm. The mobile phase consisted of Milli-Q water and methanol containing 0.05% perchloric acid and 0.1% tetrabutyl N-ammonium hydroxide. The retention time of ETH was 4.9 minutes (12).

**Calculation of Pharmacokinetic variables:** Based on plasma concentration of drugs at different time-points, certain PK variables such as peak concentration (C_max_), time at which C_max_ was attained (T_max_), area under the concentration-time curve (AUC_0-12_) and half-life (t_1/2_) were calculated based on non-compartmental analysis using STATA 15.0 (StataCorp, College Station, Texas, USA).

### Follow-up during treatment

All patients continued to receive ATT according to NTEP guidelines. Culture results at end of intensive phase, wherever available were recorded from the treatment card of patients. Based on the culture status at end of IP, patients were divided into two groups - (i) those who were culture negative (responders) and (ii) those who remained culture positive, those who had died during IP, and those who required change in regimen (non-responders).

**Statistical Evaluation:** Data were analysed using STATA 15.0 (StataCorp, College Station, Texas, USA). Shapiro-Wilks test was used to assess normality of the PK data. Values were expressed as median and range. Non-parametric Mann-Whitney U test was used to compare subgroups. Proportion of patients having C_max_ within the therapeutic ranges (8 - 13μg/ml for LFX; 20 - 60μg/ml for PZA; 2 - 5μg/ml for ETH; 20 - 35μg/ml for CS; 3 - 6μg/ml for INH 300/600mg; 9 - 15μg/ml for INH 900mg) (13) were calculated. Drug C_max_ and AUC_0-12_ were compared between responders and non-responders to ATT. Multiple linear regression analysis by stepwise method was carried out to identify factors that influenced C_max_ and AUC_0-8_ of drugs. Logistic regression model was used to identity the association of C_max_ and AUC_0-12_ with culture negativity at the end of IP. Some of the following factors such as age, gender, body weight, smoking status, alcoholism, DM, culture and drug doses were considered in the regression model after considering the variance inflation factor (VIF) and co-linearity. A p ≤ 0.05 was considered statistically significant.

### Sample Size

The sample size was calculated based on the study of Mpagama et al (6), who reported the peak concentration of ETH to be 3.6μg/ml with the standard deviation of 1.8. Assuming a marginal error of 20% and 10% refusal for blood draw, the sample size required was 346 patients.

## Results

There were two groups of patients; those who were initially recruited received the LFX containing regimen (Regimen 1) and subsequently when the programme revised the regimen, the patients were being treated with the MFX-containing regimen (Regimen 2). A total of 350 patients took part in the study, among whom 274 and 76 patients respectively received regimens 1 and 2. We analysed combined data from both regimens and also performed regimen-wise analysis. We present results obtained from the combined data. Details of all the patients recruited to the study are shown in Table 2. Overall, patients with DM constituted 42% of the study population. About 62.3% of the patients had BMI <18.5kg/sq.m. A high proportion of patients were males (82.3%), had pulmonary TB (97.1%) and were previously treatment with Category II ATT regimen (70.3%).

**Table 2:**
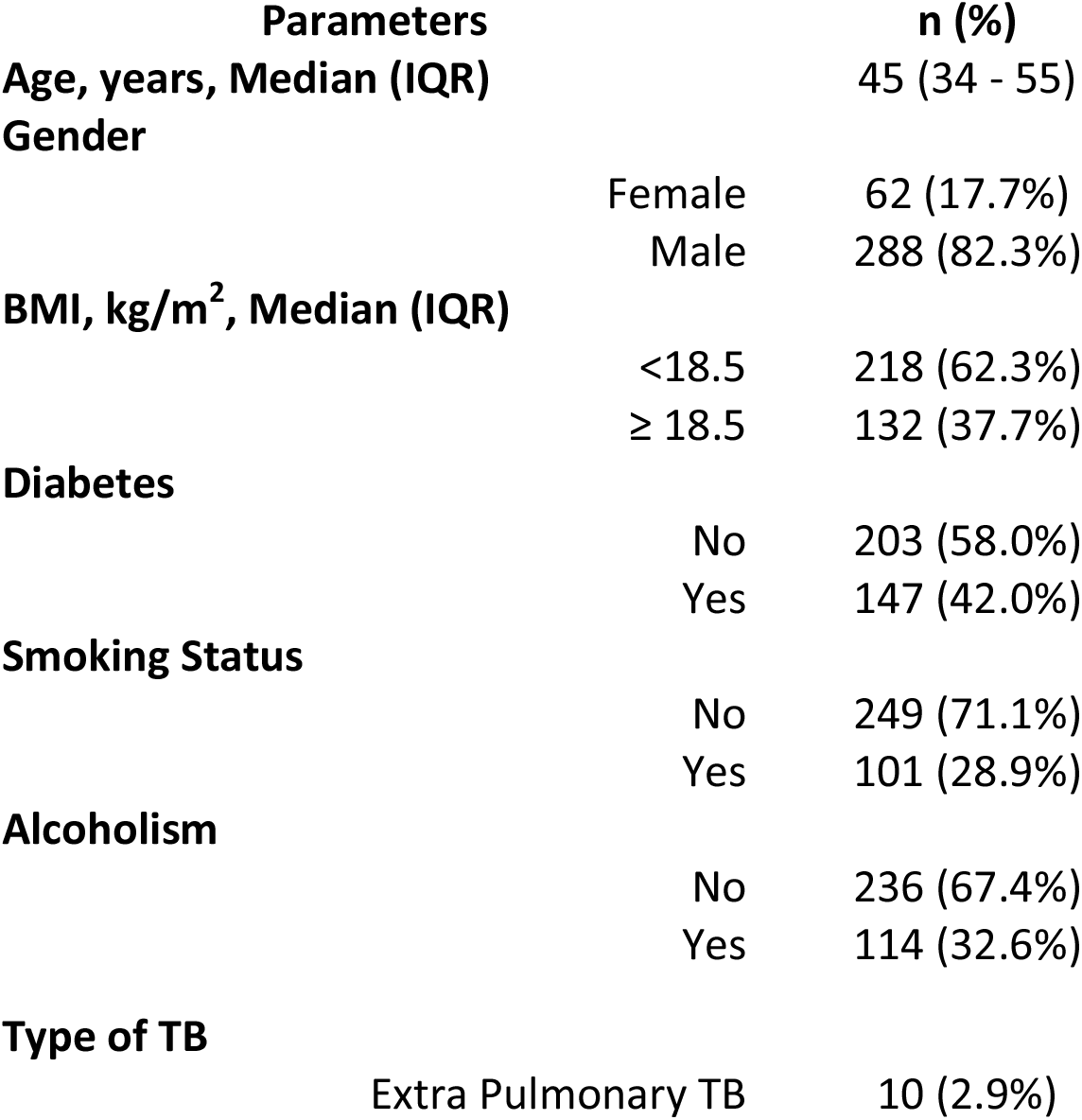
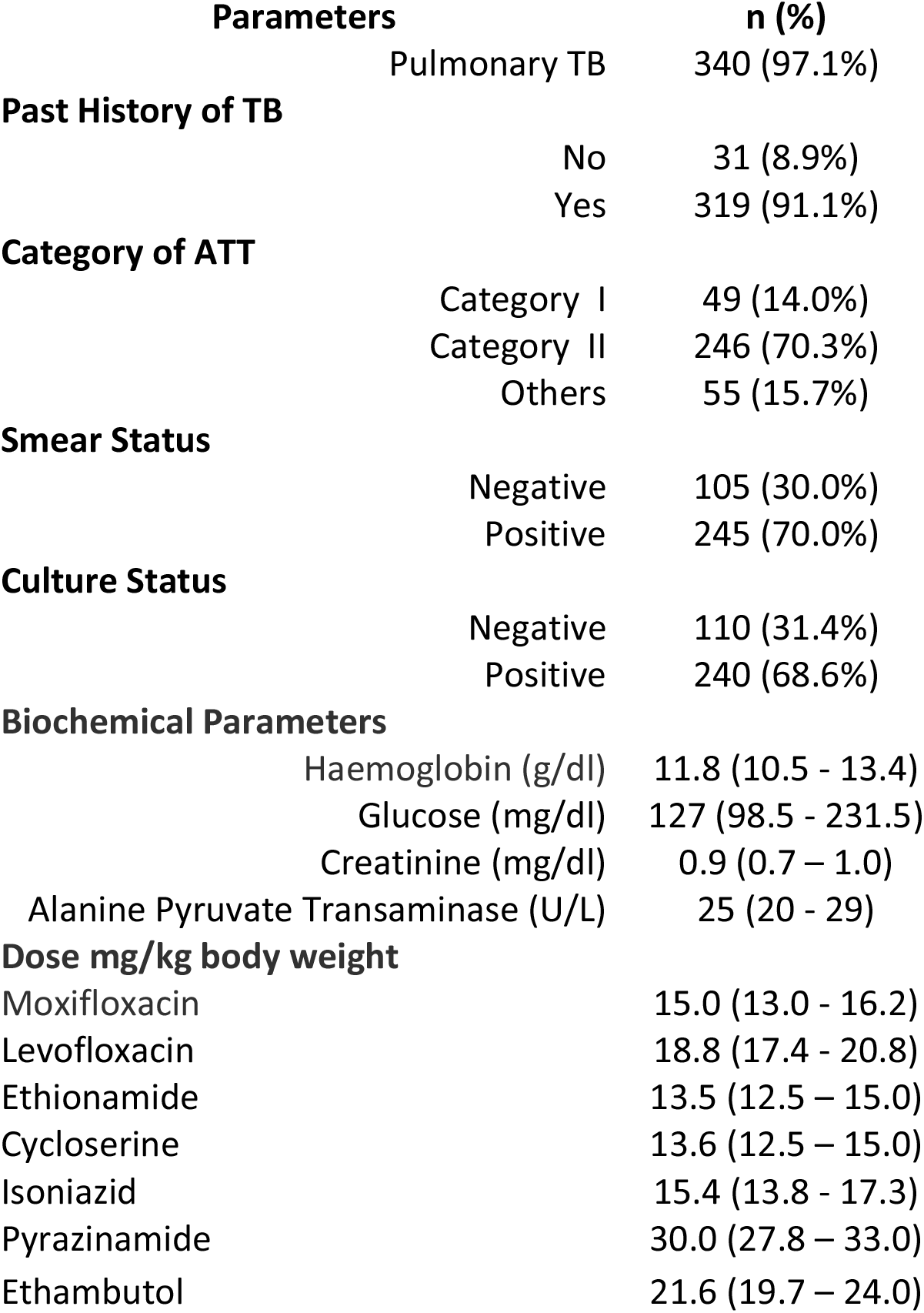
Patient Details (n = 350)

The PK parameters of LFX, ETH, CS, PZA, MFX, and INH are shown in Table 3. PK data was available for 252, 259, 235, 300, 69, 63 patients for LFX, ETH, CS, PZA, MFX and INH respectively. Missing values were mainly because patient had not taken a particular drug on the study day or blood sample was not sufficient to undertake drug estimation. The median T_max_ was 2 hours for all the drugs tested in this study. The number of patients with C_max_ of LFX within the therapeutic range (8 - 13μg/ml) was 126 (50%). The corresponding numbers for ETH (2 - 5μg/ml), CS (20 - 35μg/ml), PZA (20 - 60μg/ml), MFX (3 - 5μg/ml), INH 300mg/600mg (3 - 6μg/ml), INH 900mg (9 - 15μg/ml) were 171 (66%), 83 (35.3%), 211 (70.3%), 22 (31.9%), 3 (7.5%) and 11 (47.8%) respectively. A high proportion of patients had C_max_ of CS (57.9%), MFX (58%) and INH 300mg/600mg (85%) above the upper limit of the therapeutic range. Those with sub-therapeutic C_max_ ranged from 3.3% for PZA to 27.4% for ETH.

**Table 3:** Pharmacokinetic parameters of drugs (Values are given as Median & Inter quartile range)

| PK Parameters | Levofloxacin<br>n = 252 | Ethionamide<br>n = 259 | Cycloserine<br>n = 235 | Pyrazinamide<br>n = 300 | Moxifloxacin<br>n = 69 | Isoniazid<br>(300/600mg dose)<br>n = 40 | Isoniazid<br>(900mg dose)<br>n = 23 |
| --- | --- | --- | --- | --- | --- | --- | --- |
| C<sub>max</sub>, µg/ml | 11.6<br>(9.3 - 14.4) | 2.5<br>(2.0 - 2.5) | 37.5<br>(28.3 - 47.9) | 48.9<br>(38.7 - 60.3) | 5.6<br>(3.9 - 8.1) | 10.5<br>(7.8 - 15.9) | 11.8<br>(7.2 - 19.5) |
| T<sub>max</sub>, h | 2<br>(1 - 4) | 2<br>(2 - 2) | 2<br>(2 - 4) | 2<br>(2 - 4) | 2<br>(2 - 4) | 2<br>(1 - 2) | 2<br>(1 - 2) |
| AUC<sub>0-12</sub>, µg/ml.h | 88.6<br>(69.7 - 111.4) | 15.2<br>(10.5 - 15.2) | 344.3<br>(263.8 - 454.6) | 453.9<br>(336.4 - 576.6) | 43.1<br>(30.1 - 64.9) | 58.3<br>(40.0 - 102.4) | 74.6<br>(36.7 - 113.1) |
| T<sub>1/2</sub>, h | 8.0<br>(6.2 - 11.2) | 3.4<br>(2.2 - 3.4) | 16.3<br>(11.6 - 24.1) | 12.7<br>(9.6 - 16.8) | 7.0<br>(5.7 - 8.8) | 4.8<br>(3.6 - 6.3) | 4.7<br>(3.5 - 6.6) |
| **No (%) of Patients with C<sub>max</sub>** |  |  |  |  |  |  |  |
| Below therapeutic Range | 34 (13.5%) | 71 (27.4%) | 16 (6.8%) | 10 (3.3%) | 7 (10.1%) | 3 (7.5%) | 6 (26.1%) |
| Within therapeutic Range | 126 (50.0%) | 171 (66.0%) | 83 (35.3%) | 211 (70.3%) | 22 (31.9%) | 3 (7.5%) | 11 (47.8%) |
| Above therapeutic Range | 92 (36.5%) | 17 (6.6%) | 136 (57.9%) | 79 (26.3%) | 40 (58.0%) | 34 (85.0%) | 6 (26.1%) |

Therapeutic Range: LFX: 8 – 13 µg/ml; ETH: 2 – 5 µg/ml; CS: 20 – 35 µg/ml; PZA: 20 – 60 µg/ml; MFX: 3 – 5 µg/ml; INH: 3 – 6 µg/ml; C<sub>max</sub> = Peak Concentration; T<sub>max</sub> = Time at which peak concentration was attained; AUC = area under the time concentration curve; T<sub>1/2</sub> = half-life

Drug C_max_ and AUC_0-12_ of the different groups of patients are shown in Tables 4 and 5 respectively. Patients above 45 years of age had significantly higher C_max_ and AUC_0-12_ of LFX than those below 45 years (C_max_: 12.0 vs 11.2μg/ml, p = 0.041; AUC_0-12_: 94.7 vs 82.0μg/ml.h, p = 0.004). The AUC_0-12_ of CS was significantly higher in male than female patients (354.8 vs 307.7, p = 0.004). The C_max_ and AUC_0-12_ of LFX and CS were significantly higher in patients with BMI ≥ 18.5 than those with BMI < 18.5. Patients with DM had significantly higher C_max_ of LFX (12.0 vs 11.1μg/ml; p = 0.014) and CS (42.4 vs 35.5μg/ml; p = 0.004) than those without DM. The AUC_0-12_ of CS was also significantly higher in those with DM than those without DM (388.7 vs 323.9μg/ml.h; p = 0.004). Patients who consumed alcohol had higher C_max_ and AUC_0-12_ of CS, the difference attaining statistical significance for AUC_0-12_ only (383.9 vs 323.4μg/ml.h; p = 0.017). Non-responding patients had significantly lower C_max_ (2.5 vs 2.9μg/ml; p = 0.040) and AUC_0-12_ (14.4 vs 16.8μg/ml.h; p = 0.034) of ETH than responders at end of IP.

**Table 4:** C_max_ of drugs among different patient groups.

| Parameters | Levofloxacin<br>(µg/ml) |  | Ethionamide<br>(µg/ml) |  | Cycloserine<br>(µg/ml) |  | Pyrazinamide<br>(µg/ml) |  | Moxifloxacin<br>(µg/ml) |  | Isoniazid<br>(µg/ml) |  |
| --- | --- | --- | --- | --- | --- | --- | --- | --- | --- | --- | --- | --- |
|  | n | Median (IQR) | n | Median (IQR) | n | Median (IQR) | n | Median (IQR) | n | Median (IQR) | n | Median (IQR) |
| Age, years |  |  |  |  |  |  |  |  |  |  |  |  |
| <45 | 128 | 11.2 (9.1 - 13.7) | 121 | 2.4 (1.9 - 3.4) | 120 | 37.0(28.3 - 45.4) | 140 | 50.6 (39.4 - 59.8) | 26 | 5.1 (3.6 - 7.4) | 25 | 9.9 (7.8 - 14.2) |
| ≥45 | 124 | 12.0 (9.6 - 15.9) | 138 | 2.7 (2.0 - 3.8) | 115 | 38.5 (28.3 - 48.7) | 160 | 47.9 (37.8 - 60.5) | 43 | 5.7 (4.1 - 8.1) | 38 | 13.1 (7.9 - 16.9) |
| P Value |  | 0.041 |  | 0.151 |  | 0.222 |  | 0.791 |  | 0.396 |  | 0.232 |
| Gender |  |  |  |  |  |  |  |  |  |  |  |  |
| Female | 38 | 11.6 (9.3 - 15.1) | 37 | 2.5 (1.9 - 3.9) | 34 | 33.6 (26.5 - 40.6) | 57 | 52.1 (41.1 - 59.6) | 20 | 5.2 (3.9 – 7.0) | 18 | 11.5 (7.5 - 16.1) |
| Male | 214 | 11.6 (9.4 - 14.3) | 222 | 2.6 (2.0 - 3.6) | 201 | 37.9 (29.2 - 48.1) | 243 | 48.6 (38.2 - 60.5) | 49 | 5.7 (3.9 - 8.1) | 45 | 11.1 (7.9 - 15.7) |
| P Value |  | 0.759 |  | >0.99 |  | 0.071 |  | 0.802 |  | 0.832 |  | 0.796 |
| BMI, kg/m<sup>2</sup> |  |  |  |  |  |  |  |  |  |  |  |  |
| <18.5 | 157 | 11.2 (8.7 - 13.7) | 151 | 2.5 (1.8 - 3.5) | 147 | 36.2 (27.5 - 46.1) | 184 | 47.9 (38.3 - 60.9) | 41 | 6.1 (4.1 - 8) | 41 | 11.1 (7.8 - 15.7) |
| ≥18.5 | 95 | 12.2 (10.9 - 15.1) | 108 | 2.6 (2.1 - 3.9) | 88 | 42.5 (31.6 - 48.4) | 116 | 50.1 (39.5 – 60.0) | 28 | 5.0 (3.5 - 8.2) | 22 | 10.7 (8.0 - 19.5) |
| P Value |  | 0.004 |  | 0.051 |  | 0.025 |  | 0.977 |  | 0.396 |  | 0.806 |
| Smoking Status |  |  |  |  |  |  |  |  |  |  |  |  |
| No | 177 | 11.6 (9.4 - 13.8) | 182 | 2.5 (2.0 - 3.6) | 167 | 37.2 (27.5 - 48.6) | 214 | 50.9 (38.2 - 60.2) | 52 | 5.5 (3.9 - 7.8) | 48 | 11 (7.6 - 16.3) |
| Yes | 75 | 11.6 (8.7 - 15.6) | 77 | 2.8 (2.0 - 3.9) | 68 | 38.9 (30.8 - 47.5) | 86 | 47.7 (39.5 - 61.3) | 17 | 5.7 (4.3 - 8.2) | 15 | 12.4 (7.9 - 14.6) |
| P Value |  | 0.814 |  | 0.611 |  | 0.406 |  | 0.960 |  | 0.723 |  | 0.987 |
| Alcoholism |  |  |  |  |  |  |  |  |  |  |  |  |
| No | 166 | 11.4 (9.4 - 13.8) | 171 | 2.5 (1.9 - 3.6) | 153 | 36.2 (27.3 - 46.1) | 200 | 50.4 (37.8 - 60.1) | 50 | 5.6 (3.9 - 8.2) | 46 | 10.9 (7.2 - 16.6) |
| Yes | 86 | 11.8 (8.9 - 15.6) | 88 | 2.9 (2.0 - 3.9) | 82 | 39.7 (31.2 - 48.3) | 100 | 48.0 (39.4 - 62.2) | 19 | 5.6 (3.8 – 8.0) | 17 | 12.4 (9.1 - 14.3) |
| P Value |  | 0.419 |  | 0.258 |  | 0.055 |  | 0.879 |  | 0.523 |  | 0.733 |
| Diabetes |  |  |  |  |  |  |  |  |  |  |  |  |
| No | 141 | 11.1 (9.0 - 13.7) | 147 | 2.5 (2.0 - 3.6) | 134 | 35.5 (27.4 - 45.1) | 172 | 50.7 (39.3 - 60.4) | 43 | 5.6 (4.0 – 8.0) | 43 | 12.4 (7.8 - 16.1) |
| Yes | 111 | 12.0 (10.0 - 15.1) | 112 | 2.6 (1.9 - 3.9) | 101 | 42.4 (31.2 - 49.4) | 128 | 48.0 (37.8 - 60.3) | 26 | 5.6 (3.9 - 8.2) | 20 | 9.6 (6.4 - 17.3) |
| P Value |  | 0.014 |  | 0.891 |  | 0.004 |  | 0.683 |  | 0.911 |  | 0.516 |
| Culture Status |  |  |  |  |  |  |  |  |  |  |  |  |
| Negative | 59 | 11.6 (9.1 - 14.6) | 84 | 2.9 (2.0 - 3.8) | 52 | 37.1 (27.2 – 46.0) | 96 | 50.1 (40.0 - 58.9) | 39 | 5.4 (3.9 - 8.2) | 35 | 13.6 (8.5 - 16.1) |
| Positive | 193 | 11.5 (9.4 - 14.3) | 175 | 2.5 (1.9 - 3.6) | 183 | 37.6 (29.2 - 48.3) | 204 | 48.7 (37.4 - 60.5) | 30 | 5.7 (4.0- 8.0) | 28 | 9.5 (6.6 - 15.8) |
| P Value |  | 0.857 |  | 0.040 |  | 0.525 |  | 0.854 |  | 0.785 |  | 0.125 |

**Table 5:** AUC_0–12_ of drugs among different patient groups.

| Parameters | Levofloxacin<br>(µg/ml) |  | Ethionamide<br>(µg/ml) |  | Cycloserine<br>(µg/ml) |  | Pyrazinamide<br>(µg/ml) |  | Moxifloxacin<br>(µg/ml) |  | Isoniazid<br>(µg/ml) |  |
| --- | --- | --- | --- | --- | --- | --- | --- | --- | --- | --- | --- | --- |
|  | n | Median (IQR) | n | Median (IQR) | n | Median (IQR) | n | Median (IQR) | n | Median (IQR) | n | Median (IQR) |
| **Age, years** |  |  |  |  |  |  |  |  |  |  |  |  |
| <45 | 128 | 82.0 (67.0 - 105.2) | 121 | 13.6 (10.0 - 20.4) | 120 | 329.6 (260.6 - 436.1) | 140 | 458.0 (336.4 - 560.3) | 26 | 38.2 (27.1 - 60.3) | 25 | 58.0 (40.6 - 83.8) |
| ≥45 | 124 | 94.7 (73.6 - 119.6) | 138 | 15.9 (11.3 - 21.8) | 115 | 353.3 (266.6 - 466.2) | 160 | 448.1 (335.4 - 605.0) | 43 | 44.7 (32.1 - 69.9) | 38 | 75.7 (39.7 - 112.4) |
| P Value |  | 0.004 |  | 0.076 |  | 0.117 |  | 0.637 |  | 0.373 |  | 0.232 |
| **Gender** |  |  |  |  |  |  |  |  |  |  |  |  |
| Female | 38 | 82.1 (61.5 - 103.8) | 37 | 13.4 (10.0 - 21.1) | 34 | 307.7 (227.1 - 392.3) | 57 | 423.2 (313.1 - 552.2) | 20 | 36.7 (30.4 - 53.0) | 18 | 66.4 (43.4 - 102.3) |
| Male | 214 | 89.9 (70.3 - 113.0) | 222 | 15.4 (11.3 - 20.8) | 201 | 354.8 (267.0 - 460.1) | 243 | 454.7 (339.1 - 594.2) | 49 | 47.9 (30.1 - 69.9) | 45 | 65.2 (39.7 - 102.4) |
| P Value |  | 0.134 |  | 0.716 |  | 0.004 |  | 0.269 |  | 0.308 |  | 0.704 |
| **BMI, kg/m<sup>2</sup>** |  |  |  |  |  |  |  |  |  |  |  |  |
| <18.5 | 157 | 85.2 (66.0 - 108.9) | 151 | 13.7 (9.9 - 20.3) | 147 | 322.7 (260.2 - 447.7) | 184 | 443.7 (335.2 - 591.3) | 41 | 49.3 (32.4 - 64.2) | 41 | 63.9 (39.7 - 97.1) |
| ≥18.5 | 95 | 94.3 (75.8 - 112.9) | 108 | 15.7 (11.7 - 21.9) | 88 | 401.6 (296.6 - 457.4) | 116 | 468.4 (336.9 - 568.9) | 28 | 36.7 (29.1 - 67.4) | 22 | 75.4 (40.6 - 113.1) |
| P Value |  | 0.048 |  | 0.060 |  | 0.015 |  | 0.629 |  | 0.406 |  | 0.395 |
| **Smoking Status** |  |  |  |  |  |  |  |  |  |  |  |  |
| No | 177 | 87.8 (69.8 - 112.9) | 182 | 15.2 (10.4 - 21.1) | 167 | 333.1 (258.2 - 454.6) | 214 | 451.5 (333.9 - 573.7) | 52 | 42.0 (29.8 - 62.3) | 48 | 64.8 (40.0 - 104.1) |
| Yes | 75 | 90.7 (68.9 - 111.4) | 77 | 15.4 (11.3 - 20.4) | 68 | 369.5 (287.7 - 453.5) | 86 | 454.9 (341.8 - 594.2) | 17 | 44.7 (32.1 - 71.3) | 15 | 65.2 (39.7 - 97.1) |
| P Value |  | 0.579 |  | 0.905 |  | 0.218 |  | 0.671 |  | 0.568 |  | 0.910 |
| **Alcoholism** |  |  |  |  |  |  |  |  |  |  |  |  |
| No | 166 | 87.5 (67.4 - 111.3) | 171 | 14.4 (10.0 - 20.8) | 153 | 323.4 (251.3 - 446.8) | 200 | 457.5 (332.2 - 574.2) | 50 | 42.0 (30.1 - 64.9) | 46 | 64.8 (36.7 - 103.8) |
| Yes | 86 | 90.8 (70.6 - 117.6) | 88 | 17.6 (11.5 - 21.4) | 82 | 383.9 (302.6 - 466.2) | 100 | 451.0 (347.0 - 580.0) | 19 | 43.5 (29.4 - 66.8) | 17 | 65.2 (40.6 - 85.2) |
| P Value |  | 0.321 |  | 0.226 |  | 0.017 |  | 0.754 |  | 0.747 |  | 0.865 |
| **Diabetes** |  |  |  |  |  |  |  |  |  |  |  |  |
| No | 141 | 84.3 (65.6 - 108.9) | 147 | 14.1 (10.8 - 21.4) | 134 | 323.9 (251.2 - 430.4) | 172 | 460.6 (339.9 - 583.4) | 43 | 43.1 (31.6 - 64.2) | 43 | 73.3 (39.7 - 102.4) |
| Yes | 111 | 93.8 (75.8 - 112.9) | 112 | 15.5 (10.4 - 20.4) | 101 | 388.7 (300.5 - 466.2) | 128 | 449.5 (333.6 - 571.6) | 26 | 42.3 (29.5 - 69.8) | 20 | 56.2 (38.7 - 107.7) |
| P Value |  | 0.183 |  | 0.989 |  | 0.004 |  | 0.635 |  | 0.862 |  | 0.690 |
| **Culture Status** |  |  |  |  |  |  |  |  |  |  |  |  |
| Negative | 59 | 87.1 (67.0 - 114.5) | 84 | 16.8 (11.4 - 23.0) | 52 | 337.6 (238.6 - 437.1) | 96 | 453.9 (336.9 - 560.6) | 39 | 41.1 (29.4 - 64.9) | 35 | 78.2 (48.3 - 104.3) |
| Positive | 193 | 89.4 (70.3 - 109.0) | 175 | 14.4 (10.0 - 20.2) | 183 | 348.8 (264.1 - 459.1) | 204 | 454.1 (335.6 - 591.9) | 30 | 43.3 (32.4 - 66.8) | 28 | 49.9 (36.8 - 82.0) |
| P Value |  | 0.933 |  | 0.034 |  | 0.250 |  | 0.933 |  | 0.654 |  | 0.638 |

In multiple linear regression model, we determined factors that had an impact on C_max_ and AUC_0-12_ of drugs. Using C_max_ as a dependent variable patients with BMI > 18.5kg/m^2^ were likely to have 1.4 and 0.3μg/ml respectively of LFX and ETH C_max_ higher than those with BMI <18.5kg/m^2^. Patients with culture positive *M. tuberculosis* were likely have 0.4μg/ml ETH C_max_ lower than those with culture negative *M. tuberculosis*. An increase of one unit of drug dose taken was likely to cause increases in the C_max_ of PZA, MFX and INH by 0.5μg/ml, 0.3μg/ml and 0.7μg/ml respectively.

Using AUC_0-12_ as the dependent variable and adjusting for co-variates, age (LFX and ETH), gender (CS), BMI (LFX, ETH and CS), alcoholism (ETH), culture status (ETH), and mg/kg drug dose (ETH, PZA and MFX) were significant.

Of the 350 patients recruited to the study, status at end of IP were available for 303 patients in the NTEP records. Among them, 214 were responders, while 45 patients were non-responders at end of IP. Patients who had defaulted treatment (n = 44) were excluded from analysis. The median C_max_ (2.0 vs 2.9μg/ml; p = 0.005) and AUC_0-12_ (12.2 vs 17.0μg/ml.h; p = 0.002) of ETH were significantly lower in non-responders compared to responders (Figures 1A and B).

**Figure 1:**
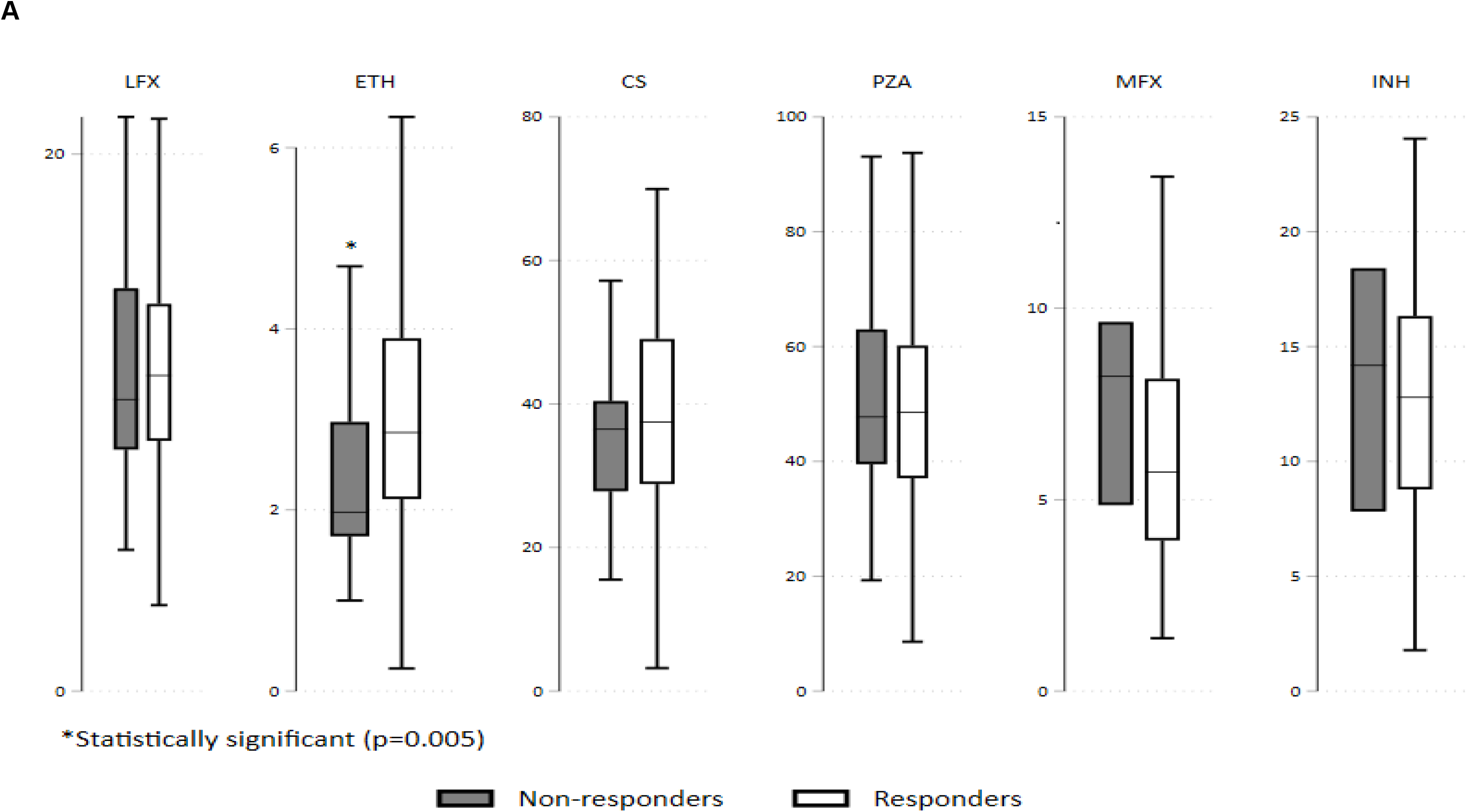
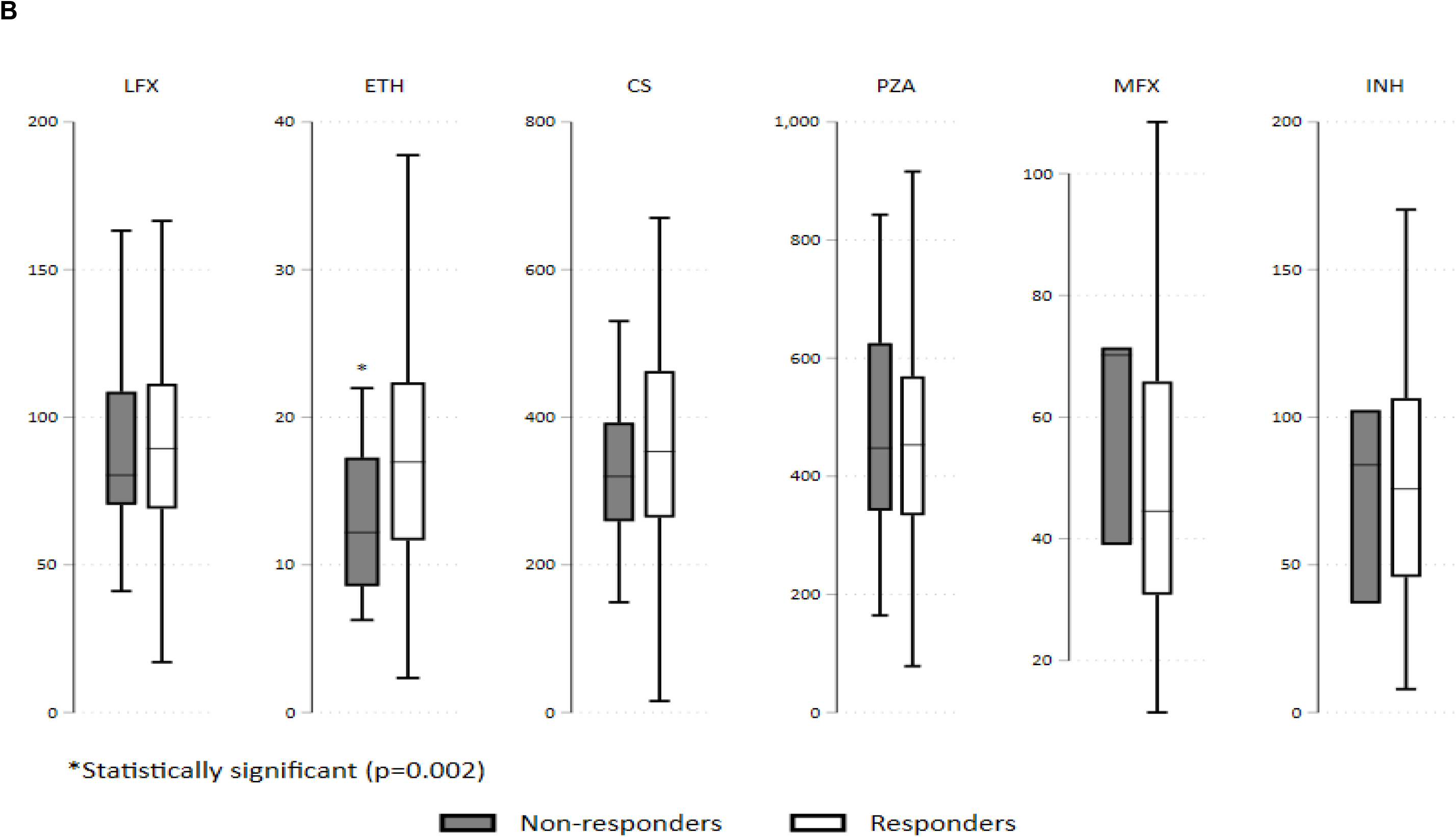
Boxplot (Median & IQR) comparing C_max_ (A) and AUC_0-12_ (B) of drugs between responders and non-responders.

The influence of factors such as age, gender, body weight, smoking, alcohol use, DM and C_max_/AUC_0-12_ of LFX, ETH, CS, PZA, MFX and INH on end of IP status were tested. After excluding defaulters and adjusting for confounders, AUC_0-12_ of ETH was observed to significantly influence end of IP status (aOR - 1.065; 95% CI: 1.001 - 1.134; p = 0.047). The chance of having culture negativity at end of IP was higher by having AUC_0-12_ of ETH increased by 7% (Table 6).

**Table 6:** Impact of drug levels on status at end of intensive phase.

| Drug | Peak Concentration |  |  |  | Exposure |  |  |  |
| --- | --- | --- | --- | --- | --- | --- | --- | --- |
|  | OR (95% CI) | P Value | aOR (95% CI) | P Value | OR (95% CI) | P Value | aOR (95% CI) | P Value |
| LFX | 0.993 | 0.872 | 1.001 | >0.950 | 0.999 | 0.802 | 0.999 | 0.875 |
|  | (0.915 - 1.078) |  | (0.919 - 1.090) |  | (0.990 - 1.008) |  | (0.990 - 1.009) |  |
| ETH | 1.321 | 0.118 | 1.262 | 0.198 | 1.068 | 0.034 | 1.065 | 0.047 |
|  | (0.931 - 1.874) |  | (0.885 - 1.799) |  | (1.005 - 1.136) |  | (1.001 - 1.134) |  |
| CS | 1.014 | 0.332 | 1.010 | 0.503 | 1.002 | 0.273 | 1.001 | 0.498 |
|  | (0.986 - 1.043) |  | (0.981 - 1.041) |  | (0.999 - 1.004) |  | (0.998 - 1.004) |  |
| PZA | 0.992 | 0.429 | 0.993 | 0.523 | 0.999 | 0.509 | 0.999 | 0.523 |
|  | (0.972 - 1.012) |  | (0.974 - 1.014) |  | (0.997 - 1.001) |  | (0.997 - 1.001) |  |
| INH | 0.974 | 0.745 | 0.960 | 0.814 | 1.000 | >0.950 | 1.043 | 0.912 |
|  | (0.828 - 1.144) |  | (0.799 - 1.240) |  | (0.974 - 1.026) |  | (0.980 - 1.040) |  |
| MFX | 0.821 | 0.318 | 0.880 | 0.451 | 0.976 | 0.333 | 0.990 | 0.512 |
|  | (0.557 - 1.210) |  | (0.520 - 1.490) |  | (0.930 - 1.025) |  | (0.910 - 1.240) |  |

Odds ratio was calculated after adjusting for the factors such as age, gender, body weight, smoking status, alcoholism and diabetes mellitus

## Discussion

Effective control of MDR TB remains a challenge since second-line anti-TB medications are less potent, may require longer treatment duration, have a narrow therapeutic range and have greater number of side effects than first-line drugs (14). Hence treatment outcomes for MDR TB remain sub-optimal compared to drug-susceptible TB. Furthermore, PK tools such as plasma drug concentrations and MIC testing are not readily available in most TB endemic settings and it remains unclear how such measurements are best utilised.

The importance of optimised drug exposure, leading to greater bacterial killing and better outcomes has been shown in both murine models and human experience (15 - 17). In this prospective cohort study, we have described the PK of anti-TB drugs used in the treatment of MDR TB in India, and factors that were likely to influence status at end of IP. A higher proportion of patients had C_max_ of LFX, ETH, PZA and INH 900mg within the therapeutic range. While this is quite encouraging, we did observe a higher proportion of patients having C_max_ of CS, MFX, and INH 300/600mg above the therapeutic range. The reason for higher proportion of patients who received a relatively lower INH dose having C_max_ above the therapeutic range than those who received a higher dose remains unclear, although patients’ INH acetylator status would have thrown some light on this issue. However, INH acetylator status was not determined in this study. Our finding of 58% of patients having C_max_ of CS above the therapeutic range is in agreement with that reported by Mpagama et al, which was 52% in their study (6). Using hollow fiber system model of TB, Deshpande et al, determined the susceptibility breakpoint of CS, and reported that drug doses required to achieve bacterial killing in patients was high, which was likely to cause toxicity (18). In this study, we did not observe major adverse events due to CS at the time of the PK sampling day. It should be pointed out that majority of the patients had their PK study conducted within 4 weeks of treatment initiation. Psychosis as a side effect due to CS generally shows up after a month of treatment; however, follow-up of patients beyond the PK study was not part of this study..

Our observation that a small proportion of patients only had sub-therapeutic C_max_ points to the fact that drug doses used in the NTEP were quite adequate. A population PK study by Chigutsa and others in South African MDR TB patients reported a high proportion of patients failed to achieve the target ofloxacin exposure and suggested that LFX or MFX would be ideal fluoroquinolones to treat MDR TB (19). According to the PMDT guidelines in India, LFX was part of the multi-drug regimen and the revised shorter regimen had MFX in the place of LFX. Our observation of half of the patients having optimal C_max_ of LFX contradicts the findings of Mpagama et al, who reported LFX concentrations were frequently lower in MDR TB patients in Tanzania (6). Using nonlinear mixed-effects modelling, a population PK study of PZA suggested 1500mg, 1750mg and 2000mg PZA doses for MDR TB patients having weight bands upto 50kg, 51-70kg and above 70kg respectively (20). The PZA doses followed in the NTEP are almost similar to that recommended in the South African study.

A direct comparison of PK data of the drugs examined in this study could be compared with that reported by others, since the Korean study was performed in healthy volunteers (7) and the Tanzanian study examined drug concentrations only at 2 hours post-dosing (6). Furthermore, these studies were conducted in small numbers, 14 healthy volunteers and 25 patients.

Patients above 45 years of age seemed to have higher C_max_ and AUC_0-12_ of LFX, probably due to slower metabolism of the drug with aging. Female patients were observed to have higher exposure of CS than their male counterparts. Although not many studies have reported gender - based differences in second-line anti-TB medications, a similar trend has been observed with respect to first-line anti-TB drugs (21 - 23). Our finding of patients with higher BMI having higher C_max_ and AUC_0-12_ of LFX and CS are not consistent with that reported with respect to first-line drugs (20). Multivariate regression analysis also seemed to show BMI having a direct relationship with drug concentrations. Likewise, higher LFX and CS concentrations observed in those with DM than those without DM is also not in line with certain reports on RMP, INH and PZA (21, 24, 25). While INH and PZA concentrations were lower in drug susceptible TB patients with DM than those without DM (25, 26), no differences were observed in the case of MDR TB patients with and without DM. It could be hypothesised that the metabolic pathways of drugs are different in patients with drug susceptible and drug resistant TB, although the possibility of drug-drug interactions cannot be ruled out. Nonetheless, this observation requires some attention and confirmation in other studies.

The study showed that non-responding patients to ATT having lower C_max_ and AUC_0-12_ of ETH than responders is quite significant. We combined three different groups of patients as non-responders, which included those who required a change of regimen. According to the NTEP, MDR TB patients showing signs of clinical deterioration and becoming morbid have their treatment changed to a bedaquiline - containing regimen. Thus, culture positives at end of IP, clinical failures and deaths before completion of IP were combined and considered as non-responders. The PMDT guidelines in India, recommends ETH as part of the MDR TB treatment regimen both initially and in the revised regimen. Using a multidose hollow fiber system model, Deshpande et al, demonstrated that ETH had a reasonable kill rate and that it was an important contributor to MDR TB treatment regimens (27). Furthermore, suboptimal ETH exposure was likely to cause efflux pump - mediated acquired drug resistance. Our study findings of C_max_ and AUC_0-12_ of ETH being higher in those who had culture conversion at end of IP than those who did not, and ETH exposure emerging as a significant factor impacting end-of-IP status (after adjusting for confounding factors and excluding defaulters) are consistent with the study of Deshpande and others (27). In the light of these findings, it is crucial to include ETH as a part of the MDR TB treatment regimen, and ensure that therapeutic concentrations of ETH are maintained.

In multivariate regression analysis, we demonstrated drug doses to have a significant influence on the plasma concentrations of ETH, PZA, MFX and INH, and there was a direct relationship. Increasing drug doses was likely to boost drug concentrations, although one needs to be cautious about occurrence of toxic effects.

Our study findings are based on data analysed from all the 350 patients. We also performed regimen-wise analysis. Of the drugs examined in this study, ETH and PZA were present in both the regimens. No striking differences were observed in the PK profile of drugs in either regimen. This observation was made with respect to patients maintaining therapeutic concentrations, group-wise comparisons and factors influencing drug PK. It should however be added that the study was not designed to examine drug-drug interactions, about which not much is known.

The strength of the study was the large sample size and understanding the association between drug concentrations and patients’ status at end of IP. The study was however, limited by the fact that we did not follow up patients till end of treatment. Hence treatment outcomes and occurrence of adverse reactions to drugs were not known. However, assessing end-of-treatment outcome in MDR TB studies is quite cumbersome due to the long duration of treatment. Determination of INH acetylator status could have provided additional information.

In summary, this is the first report describing the PK of second-line anti-TB drugs in MDR TB patients in India. All patients were being treated according to the NTEP guidelines under direct supervision, ensuring treatment regularity. The study conducted in a fairly large, adequately powered sample size has demonstrated that the drug doses used currently in the programme produced optimal drug concentrations in majority of patients. ETH played a major role in the MDR TB combination regimen and was a key determinant of end-of-IP status. Future studies should adopt a comprehensive approach assessing drug exposure, individual minimum inhibitory concentrations and outcome. Researchers from China are aiming to conduct a translational study that would characterise second-line anti-TB drug exposures and relate them to individual MICs (28). It is important to carry out similar PK/PD studies in different parts of India, in order to generalise the findings.

## Data Availability

All study data is available at the institute where the research study was undertaken (National Institute for Research in Tuberculosis).

## Acknowledgements

The authors thank the patients who took part in the study, A Vijayakumar for drug estimations by HPLC, clinic nurses of NIRT for blood collections, and secretarial assistance by S. Sasikumar.

## Authors’ contributions

AKH and GR designed the study, AKH wrote the study protocol and obtained regulatory approvals, RS, PLN, SRK, DN and SK supervised patient recruitment, PLN and TB conducted the study, AKH supervised drug estimations, VS and TB performed drug estimations, NSG and AB supervised bacteriological investigations, TK performed statistical analysis and GR drafted the manuscript.

### Potential conflict of interest

None

## Notes

### Competing Interest Statement

The authors have declared no competing interest.

### Funding Statement

No extra mural funding was received to conduct this study

### Author Declarations

National Institute for Research in Tuberculosis - Institutional Ethics Committee

## References

1. Global TB report, 2019. Available at: http://www.who.int/tb/publications/global_report/en/

2. RNTCP Guidelines for Programmatic Management of Drug Resistant Tuberculosis (PMDT) in India, 2012. Available at: https://tbcindia.gov.in

3. Thomas A, Ramachandran R, Rehman F, Jaggarajamma K, Santha T, Selvakumar N, Krishnan N, Nalini SM, Sundaram V, Fraser W, Narayanan PR. Management of multi-drug resistant tuberculosis in the field - Tuberculosis Research Centre experience. Indian J Tub 2007; 54: 117–124

4. Peloquin CA. Therapeutic drug monitoring in the treatment of tuberculosis. Drugs 2002; 62: 2169–2183

5. Chang MJ, Jin B, Chae J, Yun H, Kim ES, Lee YJ, Cho YJ, Yoon HI, Lee CT, Park KU, Song J, Lee JH, Park JS. Population pharmacokinetics of moxifloxacin, cycloserine, p-aminosalicylic acid and kanamycin for the treatment of multi-drug-resistant tuberculosis. Int J Antimicrob Agents 2017; 49: 677–687. doi: 10.1016/j.ijantimicag.2017.01.024

6. Mpagama SG, Ndusilo N, Stroup S, Kumburu H, Peloquin CA, Gratz J, Houpt ER, Kibiki GS, Heyself SK. Plasma drug acitivity in patients on treatment for multi-drug resistant tuberculosis. Antimicrob Agents Chemother. 2014;58:782- 788. doi: 10.1128/AAC.01549-13

7. Park S, Oh J, Jang K, Yoon J, Moon SJ, Park SJ, Lee JH, Song J, Jang IJ, Yu KS, Chung JY. Pharmacokinetics of second-line antituberculosis drugs after multiple administrations in healthy volunteers. Antimicrob Agents Chemother 2015; 59: 4429—4435. doi: 10.1128/AAC.00354-15

8. Hemanth Kumar AK, Sudha V, Ramachandran G. A Simple and Rapid Liquid Chromatography method for determination of Levofloxacin in Plasma. SAARC J TB, Lung Diseases & HIV/AIDS, 2016; XIII: 28–33

9. Hemanth Kumar AK, Geetha Ramachandran. Simple and rapid liquid chromatography method for determination of moxifloxacin in plasma. J Chromatogr B Analyt Technol Biomed Life Sci, 2009; 877: 1205-1208. doi: 10.1016/j.jchromb.2009.02.042

10. Hemanth Kumar AK, Sudha V, Geetha Ramachandran. Simple and rapid liquid chromatography method for simultaneous determination of isoniazid and pyrazinamide in plasma. SAARC J TB, Lung diseases & HIV/AIDS 2012; 9: 13—18. DOI: https://doi.org/10.3126/saarctb.v9i1.6960

11. Hemanth Kumar AK, Polisetty AK, Sudha V, Vijayakumar A, Geetha Ramachandran. A Selective and sensitive high performance liquid chromatography assay for the determination of cycloserine in human plasma. Ind J Tub 2018; 65: 118—123. doi: 10.1128/AAC.02410-17

12. Hemanth Kumar AK, Sudha V, Geetha Ramachandran. Simple and rapid high pressure liquid chromatography methods for estimation of ethionamide in plasma. Asian J Biomed Pharmaceut Sci 2014; 04: 1 - 5

13. Abdullah A, Peloquin CA. Therapeutic drug monitoring in the treatment of tuberculosis: an update. Drugs 2014; 74: 839–854. doi: 10.1007/s40265-014-0222-8

14. Shenoi S, Heysell SK, Moll A, Friedland G. Multidrug-resistant and extensively drug-resistant tuberculosis: consequences for the global HIV community. Curr Opin Infect Dis 2009; 22:11-17. 10.1097/QCO.0b013e3283210020. doi:10.1097/QCO.0b013e3283210020

15. Zhang M, Li SY, Rosenthal IM, Almeida DV, Ahmad Z, Converse PJ, Peloquin CA, Neurmberger EL, Grosset JH. Treatment of tuberculosis with rifamycin containing regimens in immune-deficient mice. Am J Respir Crit Care Med 2011; 183:1254—1261. 10.1164/rccm.201012-1949OC

16. Chang KC, Leung CC, Yew WW, Chan SL, Tam CM. Dosing schedules of 6-month regimens and relapse for pulmonary tuberculosis. Am J Respir Crit Care Med 2006; 174:1153—1158. 10.1164/rccm.200605-637OC

17. Egelund EF, Peloquin CA. Pharmacokinetic variability and tuberculosis treatment outcomes, including acquired drug resistance. Clin Infect Dis 2012; 55:178–179. 10.1093/cid/cis366

18. Devyani D, Alffenaar JW, Kose CU, Dheda K, Chapagain ML, Simbar N, Schon T, Sturkenboom MGG, Mcllleron H, Lee PS, Koeuth T, Mpagama SG, Banu S, Foongladda S, Ogarkov O, Pholwat S, Houpt ER, Heysell SK, Gumbo T. D-Cycloserine Pharmacokinetics/Pharmacodynamics susceptibility and dosing implications in multidrug-resistant tuberculosis: A Faustian deal. Clin Infect Dis 2018; 67: S308–316. doi: 10.1093/cid/ciy624

19. Chigutsa E, Meredith S, Wiesner L, Padayatchi N, Harding J, Moodley P, Mac Kenzie WR, Weiner M, Mcllleron H, Kirkpatrick CMJ. Population pharmacokinetics of ofloxacin in South African patients with multi-drug resistant tuberculosis. Antimicrob Agents Chemother 2012; 56: 3857-3863. doi: 10.1128/AAC.00048-12

20. Chirehwa MT, McIlleron H, Rustomjee R, Mthiyane T, Onyebujoh P, Smith P, Denti P. Pharmacokinetics of pyrazinamide and optimal dosing regimens for drug-sensitive and resistant tuberculosis. Antimicrob Agents Chemother. 2017;61:1–11. doi: 10.1128/AAC.00490-17

21. A K Hemanth Kumar, T Kannan, V Chandrasekaran, V Sudha, A Vijayakumar, K Ramesh, J Lavanya, Soumya Swaminathan, Geetha Ramachandran. Pharmacokinetics of thrice weekly rifampicin, isoniazid and pyrazinamide in adult tuberculosis patients in India. Int J Tuberc Lung Dis 2016; 20:1236–1241. doi: 10.5588/ijtld.16.0048

22. Mcllleron H, Wash P, Burger A, Norman J, Folb PI, Smith P. Determinants of rifampin, isoniazid, pyrazinamide and ethambutol pharmacokinetics in a cohort of tuberculosis patients. Antimicrob Agents Chemother 2006; 50: 1170—1177

23. Ray J, Gardiner I, Marriott D. Managing antituberculosis drug therapy by therapeutic drug monitoring of rifampicin and isoniazid. Intern Med J 2003; 33:229—234

24. Nijland HM, Ruslami JR, Stalenhoef JE, Nelwan EJ, Alisjahbana B, Nelwan RHH, van der Ven JAM, Danusantoso H, Aarnoutse RE, van Crevel R. Exposure of Rifampin is strongly reduced in tuberculosis patients with type 2 diabetes. Clin Infect Dis 2006; 43: 848—854

25. Hemanth Kumar AK, Chandrasekaran V, Kannan T, Lakshmi Murali, Lavanya J, Sudha V, Soumya Swaminathan, Geetha Ramachandran. Anti-tuberculosis drug concentrations in tuberculosis patients with and without diabetes mellitus. Eur J Clin Pharmacol 2017; 73: 65—70. doi: 10.1007/s00228-016-2132-z

26. Babalik A, Ulus IH, Bakirci N, Kuyucu T, Arpag H, Dagyildizi L, Capaner E. Plasma Concentrations of Isoniazid and Rifampin are decreased in adult pulmonary tuberculosis patients with diabetes mellitus. Antimicrob Agents Chemother. 2013; 5740—5742. doi: 10.1128/AAC.01345-13

27. Deshpande D, Pasipanodya JG, Mpagama SG, Srivastava S, Bendet P, Koeuth T, Lee PS, Heysell SK, Gumbo T. Ethionamide Pharmacokinetics/Pharmacodynamics- derived dose, the role of MICs in clinical outcome, and the resistance arrow of time in multidrug-resistant tuberculosis. Clin Infect Dis 2018; 67:S317–326. doi: 10.1093/cid/ciy609

28. Forsman LD, Niward K, Hu Y, Zheng R, Zheng X, Ke R, Cai W, Hong C, Li Y, Gao Y, Werngren J, Paues J, Kuhlin J, Simonsson USH, Eliasson E, Alffenaar JW, Mansjo M, Hoffner S, Xu B, Schon T, Brushfeld J. Plasma concentrations of second-line anti-tuberculosis drugs in relation to minimum inhibitory concentrations in multidrug-resistant tuberculosis patients in China: a study protocol of a prospective observational cohort study. BMJ Open 2018; 4;8:e023899. doi: 10.1136/bmjopen-2018-023899

